# Projecting the seasonality of endemic COVID-19

**DOI:** 10.1101/2022.01.26.22269905

**Authors:** Jeffrey P. Townsend, April D. Lamb, Hayley B. Hassler, Pratha Sah, Aia Alvarez Nishio, Cameron Nguyen, Alexandra D. Tew, Alison P. Galvani, Alex Dornburg

## Abstract

**Importance:** Successive waves of infection by SARS-CoV-2 have left little doubt that COVID-19 will transition to an endemic disease, yet the future seasonality of COVID-19 remains one of its most consequential unknowns. Foreknowledge of spatiotemporal surges would have immediate and long-term consequences for medical and public health decision-making.

**Objective:** To estimate the impending endemic seasonality of COVID-19 in temperate population centers via a phylogenetic ancestral and descendent states approach that leverages long-term data on the incidence of circulating coronaviruses.

**Design:** We performed a comparative evolutionary analysis on literature-based monthly verified cases of HCoV-NL63, HCoV-229E, HCoV-HKU1, and HCoV-OC43 infection within populations across the Northern Hemisphere. Ancestral and descendent states analyses on human-infecting coronaviruses provided projections of the impending seasonality of endemic COVID-19.

**Setting:** Quantitative projections of the endemic seasonality of COVID-19 were based on human endemic coronavirus infection incidence data from New York City (USA); Denver (USA); Tampere (Finland); Trøndelag (Norway); Gothenburg (Sweden); Stockholm (Sweden); Amsterdam (Netherlands); Beijing (China); South Korea (Nationwide); Yamagata (Japan); Hong Kong; Nakon Si Thammarat (Thailand); Guangzhou (China); and Sarlahi (Nepal).

**Main Outcome(s) and Measure(s):** The primary projection was the monthly relative frequency of SARS-CoV-2 infections in each geographic locale. Four secondary outcomes consisted of empirical monthly relative frequencies of the endemic human-infecting coronaviruses HCoV-NL63, -229E, -HKU1, and -OC43.

**Results:** We project asynchronous surges of SARS-CoV-2 across locales in the Northern Hemisphere. In New York City, SARS-CoV-2 incidence is projected in late fall and winter months (Nov.–Jan.), In Tampere, Finland; Yamagata, Japan; and Sarlahi, Nepal incidence peaks in February. Gothenburg and Stockholm in Sweden reach peak incidence between November and February. Guangzhou, China; and South Korea. In Denver, incidence peaks in early Spring (Mar.). In Amsterdam, incidence rises in late fall (Dec.), and declines in late spring (Apr.). In Hong Kong, the projected apex of infection is in late fall (Nov.–Dec.), yet variation in incidence is muted across other seasons. Seasonal projections for Nakhon Si Thammarat, Thailand and for Beijing, China are muted compared to other locations.

**Conclusions and Relevance:** This knowledge of likely spatiotemporal surges of COVID-19 is fundamental to medical preparedness and expansions of public health interventions that anticipate the impending endemicity of this disease and mitigate COVID-19 transmission. These results provide crucial guidance for adaptive public health responses to this disease, and are vital to the long-term mitigation of COVID-19 transmission.

**Key Points:** 

**Question:** Under endemic conditions, what are the projected spatiotemporal seasonal surges of COVID-19?

**Findings:** We applied a phylogenetic ancestral and descendent states approach, leveraging long-term data on the incidence of circulating coronaviruses. We found that seasonal surges are expected in or near the winter months; dependent on the specific population center, infections are forecasted to surge in the late fall, winter, or early spring.

**Meaning:** Globally, endemic COVID-19 surges should be expected to occur asynchronously, often coincident with local expected surges of other human-infecting respiratory viruses.

## Introduction

The current COVID-19 pandemic has resulted in over 6.5 million deaths worldwide ^1^.

Public health interventions—especially the closing of schools, universities and banning of large gatherings—were highly effective at reducing transmission at the advent of the pandemic ^2^.

Widespread vaccination further altered the course of the pandemic, saving tens of millions of lives globally in the first year alone ^3^. However, governmental interventions are ebbing internationally. Sustained transmission is predicted to continue into the foreseeable future ^4,5^, and there is now little doubt that COVID-19 is transitioning into a global endemic disease ^6^. This impending endemicity entails continued surges of infections causing morbidity and mortality that can be mitigated with advance preparation, especially via anticipation of future seasonal infection patterns.

COVID-19 case numbers have fluctuated in different regions and at different times during the last year. However, three challenges that have faced attempts to directly estimate future seasonal infection patterns from COVID-19 data: the global variability in public health measures, evolving pandemic transmission dynamics, and the short duration since SARS-CoV-2 emergence. These factors present too many confounders to yield highly informative studies correlating infection with environmental parameters across locations ^7 7^ such as UV light, humidity, precipitation, and temperature ^8–12^. Without well-estimated correlative associations, parameters for epidemiological modeling studies are lacking. With only a few years of infection data collected in the contexts of highly volatile and heterogeneous interventions, there are qualitative inferences but little year-to-year data that can be appropriately applied to determine the seasonality of the virus ^13^. The absence of annual SARS-CoV-2 infection data without pandemic transmission dynamics or public health interventions has hampered efforts to determine COVID-19 seasonality and resulted in contradictory estimates of seasonal trends ^14–17^, and necessitates application of alternate approaches.

One approach to predicting SARS-CoV-2 seasonality relies on comparison to other endemic viruses that follow a similar route of respiratory transmission. These “flu and cold” viruses follow known seasonal patterns of infection that vary across the globe (cites) that offer a possible analogy to the annual variation expected for SARS-CoV-2 ^17–21^. However, diverse respiratory viruses exhibit divergent patterns of seasonality. For example, rhinovirus infections occur relatively frequently in April through November, compared to respiratory syncytial virus infections, which are relatively more frequent in December, January, and February ^22^. At low evolutionary divergences, there is also variance in seasonal incidence patterns: across locales in Sweden, coronavirus infection by HCoV-OC43 occurs at its highest frequency in December and January, while infections by HCoV-NL63 generally peak in February ^22,23^. This variation is a consequence of evolutionary processes.

The most powerful predictions of the impending endemic seasonality of SARS-CoV-2 will leverage precisely estimated evolutionary divergences between human-infecting coronaviruses, accumulated knowledge of seasonal HCoV coronavirus incidence, and advances in phylogenetic comparative methods that enable the unknown seasonality of SARS-CoV-2 to be estimated. We apply such an approach to the estimation of the seasonality of SARS-CoV-2 infection based on extensive long-term incidence of other coronaviruses (HCoV-OC43, HCoV-NL63, HCoV-HKU1, HCoV-229E) across major population centers. This analysis provides an unconfounded means for estimation of the seasonal force of infection that is not dependent on isolation of interventions or identification of underlying mechanisms. Our resulting projections of endemic SARS-CoV-2 seasonality provide insight into optimal long-term public policies that can be applied to high-risk periods and to the preparation of healthcare providers for temporally and spatially localized surges.

## Materials and Methods

### Study Design

We conducted a literature search to identify data on monthly verified cases of HCoV-NL63, HCoV-229E, HCoV-HKU1, and HCoV-OC43 infection within populations across the globe. To infer seasonality of SARS-CoV-2, we applied ancestral and descendent states analyses on reconstructions of the evolutionary history of human-infecting coronaviruses to estimate the expected annual changes in cases at different geographic locales. These analyses provide a global-scale projection of the likely global changes of endemic seasonality for SARS-CoV-2.

### Data acquisition

Phylogenetic tree topologies—Phylogenetic relationships of SARS-CoV-2 and the endemic human-infecting coronaviruses were based on data from 58 Alphacoronavirus, 105 Betacoronavirus, 11 Deltacoronavirus, and three Gammacoronavirus as analyzed in Townsend et al. ^24^. These estimates of the phylogenetic topology were consistent with previous hypotheses of evolutionary relationships among coronaviruses ^25–29^ and were congruent across multiple methods of inference with strong (100% bootstrap) support for all nodes. Tree topologies were inferred by multiple maximum-likelihood (ML) analyses of the concatenated DNA sequence alignment, and results were robust to alternative phylogenetic likelihood search algorithms—IQ-TREE v2.0.6 ^30^ and RAxML v7.2.8 ^31^—and to branch-length differences arising from different approaches to divergence time estimation—IQ-TREE v2.0.6 ^30^, Relative Times (RelTime; ^32^) in MEGA X v10.1.9 ^33^ and TreeTime v0.7.6 ^34^—and to a potential history of recombination among or within genes, through phylogenetic analyses using an alignment of the putative non-recombining blocks ^35^. All trees from Townsend et al ^24^ were pruned of SARS-CoV-1 and MERS-CoV branches because temporal trends of infection by these viruses reflect short-term outbreaks and not seasonal endemic circulation.

Seasonal infection data—We conducted a literature search using the PubMed and Google Scholar databases searching for terms related to coronavirus, seasonality, and the known seasonalendemic human-infecting coronaviruses (HCoV-NL63, HCoV-229E, HCoV-HKU1, and HCoV-OC43). Searches were conducted in English between October 2020–August 2021, using the names of each coronavirus lineage as a key term in addition to all combinations of: coronavirus, seasonality, environmental, incidence, infection, prevalence, latitude, temperature, humidity, weather, global, cases—with no language restrictions imposed. Seasonal infection data were extracted from published, peer-reviewed research papers that reported monthly or finer seasonal case data for three or more coronaviruses, spanning at least one year.

### Estimating the seasonality of SARS-CoV-2

To estimate the seasonality of infections by SARS-CoV-2, we first extracted the average numbers of cases per month testing positive for HCoV-NL63, -229E, -HKU1, and -OC43 for each location. We scaled these case counts by the annual total to yield proportions of the cases sampled in each month. We then performed a phylogenetically informed ancestral and descendent states analysis, executing Rphylopars v0.2.12 ^36^ on the monthly proportions of cases to estimate the proportion of yearly infection by SARS-CoV-2 each month for each location, executing Rphylopars v0.2.12 ^36^ on the monthly proportions of cases. This approach takes known trait values (here, monthly proportions of cases for endemic coronaviruses) and applies a Brownian model of trait evolution and a phylogeny to estimate unobserved trait values for a taxon or taxa, providing best linear unbiased predictions that are mathematically equivalent to universal kriging (Gaussian process regression). Phylogenetic ancestral and descendent analyses were repeated across all topologies resulting from different inference approaches (molecular trees, relative phylogenetic chronograms, and non-recombinant alignment) to assess the impact of phylogenetic inference methods on our estimation of seasonality.

## Results

Our systematic review regarding seasonal patterns of endemic coronavirus incidence identified 14 studies that met the criteria of providing at least one year of data on at least three circulating human-infecting coronaviruses within a locale. These studies spanned three continents across the Northern Hemisphere (**Table 1**). In temperate regions, endemic coronaviruses typically exhibited pronounced seasonality (**Supplementary Figs. S1–S4**).

**Table 1.**
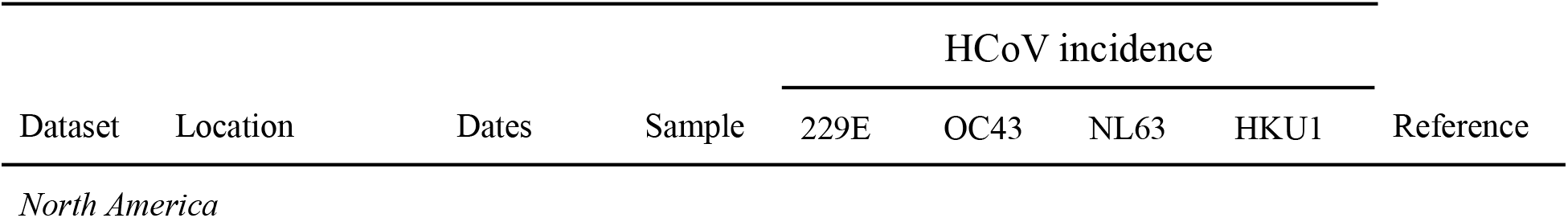
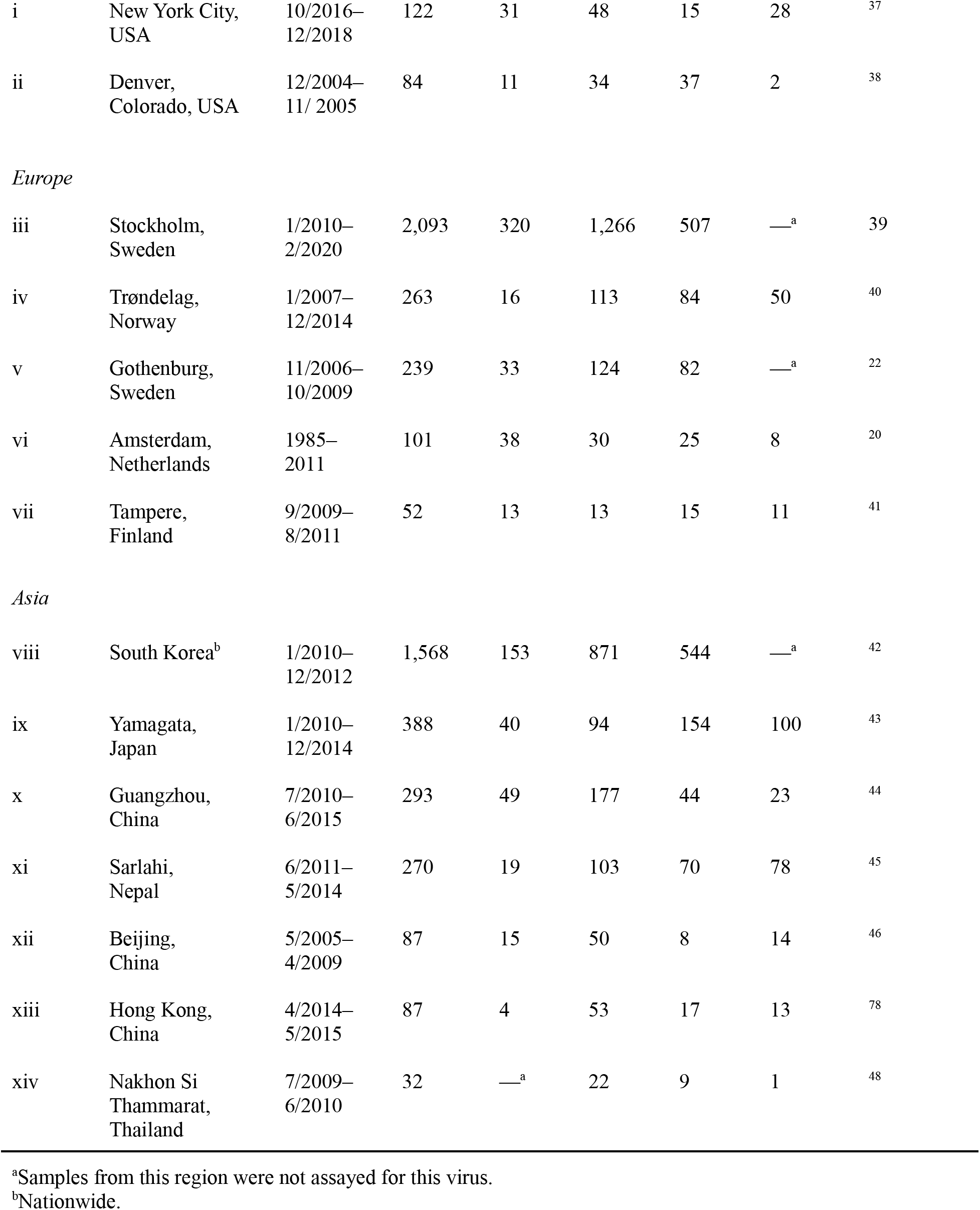
Datasets on seasonal coronavirus incidence

From our literature review, we obtained two datasets pertaining to North America. Dataset i was composed of 4215 samples taken from 196 individuals in New York City from October 2016 through April 2018 including children, teenagers, and adults with and without daily contact with children ^37^. To be included in the dataset, cases must have provided nasopharyngeal samples weekly from both nostrils for a minimum of 6 weeks. Dataset ii contained results from 1683 nasopharyngeal washes between December 2004 and November 2005 from The Children’s Hospital in Denver that tested negative for all other infectious viruses other than the viruses of interest as part of an ongoing study focusing on viral respiratory infections. Cases generally had a median age of 15 months, with a history of good health, and presented with symptoms including fever, cough, rhinorrhea, and congestion ^38^.

We obtained five datasets pertaining to Europe. Dataset iii was composed of 2,084 cases found to be positive for one of the coronaviruses, collected between 1 January 2010 and 31 December 2019 at the Karolinska University Hospital in Stockholm, Sweden ^39^. Dataset iv was composed of samples collected at St Olavs Hospital in Trondheim, Norway from children under 16 years of age that were exhibiting no symptoms and presenting for elective surgery, or that were presenting with symptoms of respiratory tract infection. ^40^. Dataset v was composed of 7,853 samples collected from November 2006 to October 2009, from 7,220 patients with a median age of 22, ranging from age 0–98 ^22^. Dataset vi was collected from serum and blood samples of adult males in the HIV-1 uninfected cohort of the Amsterdam Cohort Studies on HIV-1 and AIDS at primarily 3-to 6-month intervals spanning a 35-year period ^20^. Dataset vii was composed of stool and nasal-swab samples collected from 955 children over 2 years of age presenting with symptoms of acute respiratory infection (545), acute gastroenteritis (172), or symptoms of both (238) in the Department of Pediatrics at Tampere University Hospital in Tampere, Finland between September 2009 and August 2011 ^41^.

We obtained six datasets pertaining to Asia. Dataset viii was collected across 36 facilities in Korea by the Korea Influenza and Respiratory Viruses Surveillance System between 2013 and 2015 via throat swabs of 36,915 patients presenting with symptoms of acute respiratory infections ^42^. Dataset ix was composed of results from throat and nasal swabs of 4,342 patients (3,092 aged ≤5 years, 767 aged 6–10, 326 aged 11–15, and 104 aged >15 years) presenting with symptoms of respiratory infection in pediatric clinics in Yamagata, Japan spanning January 2010 to December 2013 ^43^. Dataset x was sourced from 13,048 throat and nasal swabs of adults and children symptomatic for acute respiratory infection between July 2010 and June 2015 in Guangzhou, China at an approximately 1.5:1 ratio of males to females ^44^. Dataset xii was composed of results from weekly nasal swabs of 3693 women enrolled in their second or third trimester of pregnancy, obtained between 2011 and 2014 in the Sarlahi district in Nepal.

Participants were enrolled in either their second or third trimester of pregnancy and were monitored until 6 months after giving birth ^45^. Dataset xiii was obtained between May 2016 to December 2017 from the bronchoalveolar lavage fluid of 408 16–90 year-old patients being treated for pneumonia or acute respiratory infection at the Central Hospital of Wuhan in Wuhan, China, with a 1.4:1 ratio of males to females among the participants ^46^. Dataset xiii was collected between April 2004 and March 2005 from 4,181 nasopharyngeal aspirates taken from patients with a mean age of 22 diagnosed with acute respiratory infection and admitted in one of two hospitals in Hong Kong: China Queen Mary Hospital and Pamela Youde Nethersole Eastern Hospital ^47^. Dataset xiv was collected from 1,254 nasopharyngeal aspirates and throat swab samples collected from patients with symptomatic acute respiratory infection in Nakhon Si Thammarat in southern Thailand between July 2009 and January 2011 ^48^.

For each location, we pruned the phylogeny of major coronavirus lineages from Townsend et al. ^24^ to include only the endemic human-infecting coronaviruses with sample data and SARS-CoV-2 (**Table 1, Fig. 1A**). To generate maximum-likelihood estimates of the spatiotemporal incidences of SARS-CoV-2, we conducted analyses of ancestral and descendent states on the relative monthly incidences for each coronavirus (**Fig. 1B–E**). All four endemic coronaviruses contributed to our projection of the relative monthly incidence of SARS-CoV-2 (**Fig. 1F**). However, the late-diverging HCoV-OC43 and HCoV-HKU1 provide more phylogenetic information than the early-diverging HCoV-NL63 and HCoV-229E. Application of this evolutionary analysis to Trøndelag, Norway provides projections that late fall and winter months will exhibit significantly higher levels of SARS-CoV-2 incidence than summer and early fall months (**Fig. 1F**).

**Figure 1.**
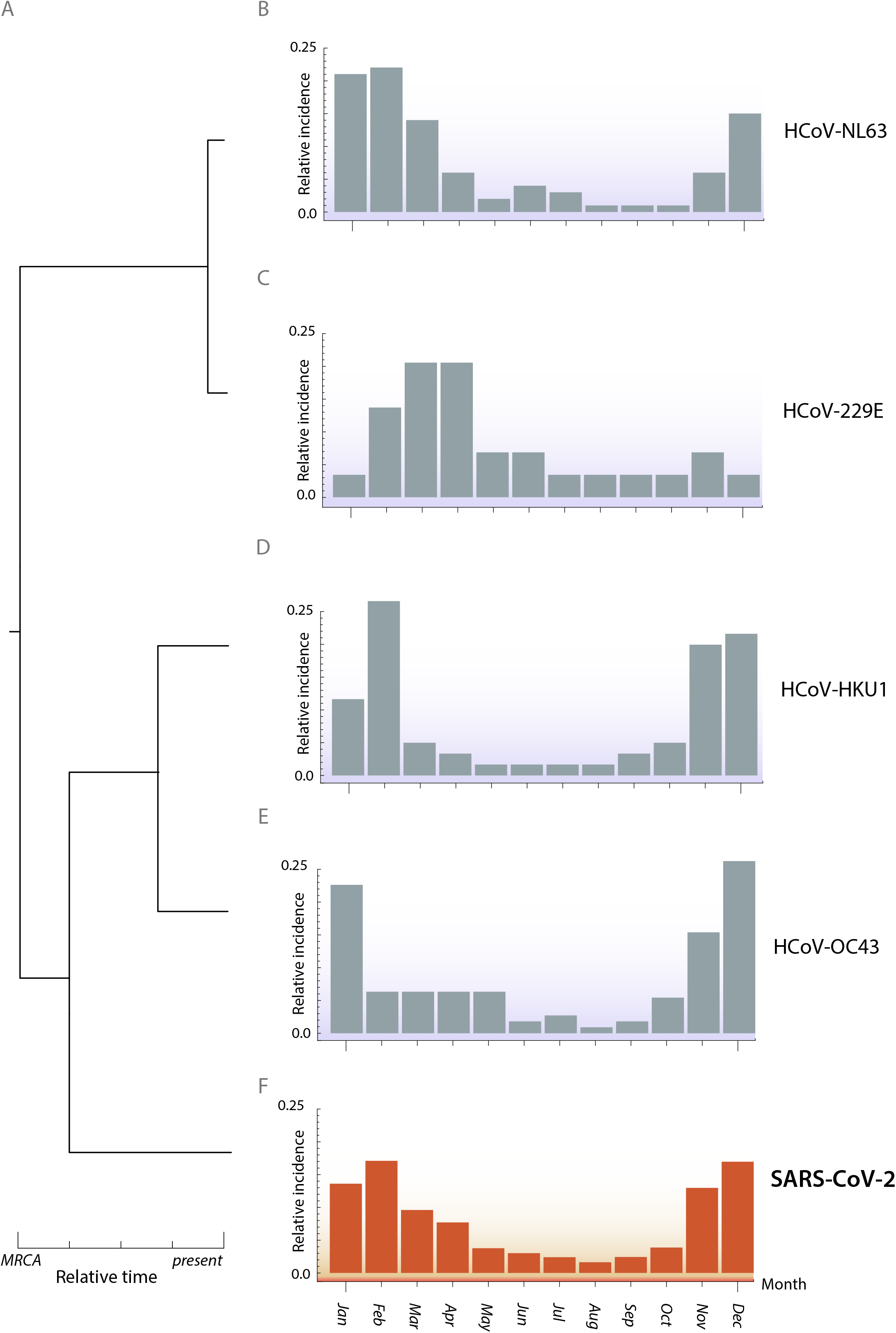
Phylogenetic inference of relative monthly incidence of SARS-CoV-2 under endemic conditions. (**A**) Time tree based on the phylogenetic divergence of circulating human-infecting coronaviruses. Empirical relative monthly incidences of HCoV (**B**) -NL63, (**C**) -229E, (**D**) -HKU1, and (**E**) -OC43, and ancestral- and descendent-states analytical estimates of relative monthly incidences of SARS-CoV-2 in Trøndelag, Norway.

This lower incidence in the summer and surrounding months is largely generalizable to much of the temperate Northern Hemisphere (**Fig. 2**). Specifically, significantly higher SARS-CoV-2 incidence is projected in late fall and winter months in New York City (**Fig. 2A**). A similar seasonality is projected for Tampere, Finland; Gothenburg and Stockholm in Sweden (**Fig. 2B**); as well as multiple locales in Asia, including Yamagata, Japan; Guangzhou, China; and South Korea (**Fig. 2C**). However, in each Northern Hemisphere continent, there are regional deviations from this seasonal pattern. In Denver, incidence is projected not to rise until the late winter, peaking in early Spring (**Fig. 2A**). Incidence in Amsterdam is similarly projected to decline in late spring. In contrast to Denver, incidence in Amsterdam rises earlier, during the late Fall (**Fig. 2B**). In Asia (**Fig. 2C**), incidence in Sarlahi, Nepal is projected to surge in early winter. In coastal, subtropical Hong Kong, the projected apex is in late fall, but monthly variation in incidence is muted across other seasons. Seasonality for tropical Nakhon Si Thammarat, Thailand is also projected to be muted relative to the temperate Northern Hemisphere locations, and the seasonality of incidence in the megalopolis of Beijing, China appears atypical with no distinct pattern. In all cases, these results were robust to the phylogenetic inference method, underlying molecular dataset, as well as the use of a chronogram or molecular evolutionary tree (**Supplementary Appendix Figs. S5–S8)**.

**Figure 2.**
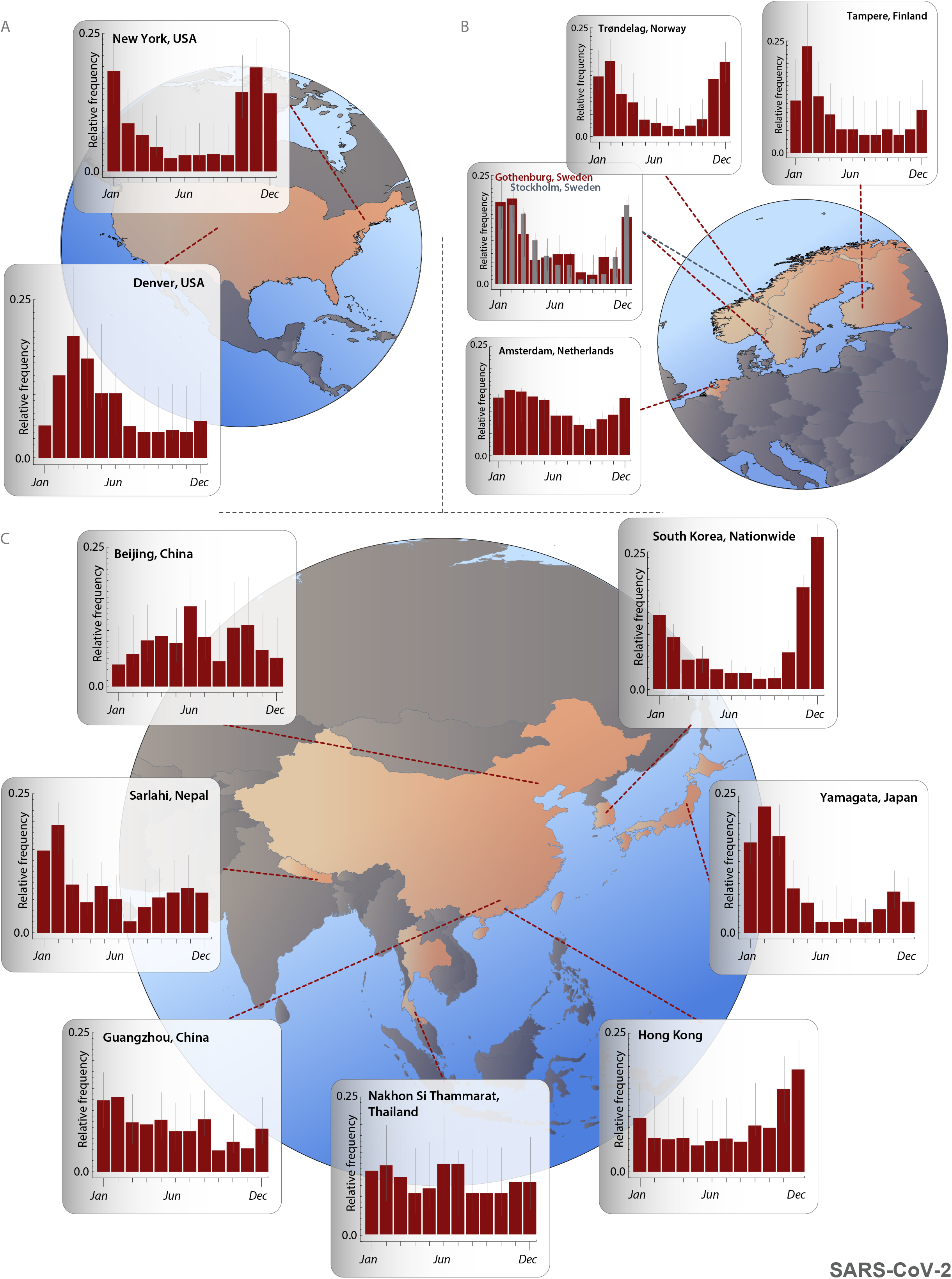
Ancestral- and descendent-states analytical estimates of the relative monthly incidence of SARS-CoV-2 under endemic conditions. (**A**) New York City and Denver, USA; (**B**) Amsterdam, Netherlands; Gothenburg and Stockholm, Sweden; Trøndelag, Norway; and Tampere, Finland; (**C**) Beijing, China; Sarlahi, Nepal; Guangzhou, China; Nakhon Si Thammarat, Thailand; Hong Kong, China; Yamagata, Japan; and South Korea (nationwide).

## Discussion

Here we analyzed monthly incidence data of the currently circulating endemic coronaviruses HCoV-NL63, -229E, -HKU1, and -OC43 to quantify seasonality of incidence of these viruses in regions that span a broad range of predominantly temperate localities across North America, Europe, and Asia. We conducted ancestral- and descendent-states analyses, projecting the seasonality of SARS-CoV-2 as it becomes endemic. Across much of the temperate Northern Hemisphere, SARS-CoV-2 can be expected to transition to a seasonal pattern of incidence that is high in late fall and winter months relative to late spring and summer. Our expected incidences through time also reveal geographic heterogeneity. This heterogeneity often manifested as a syncopation of the general northern hemispheric trend—a delay in rise to peak incidence, or a prolonged duration of higher levels of incidence relative to other areas. These temporal transmission patterns of SARS-CoV-2 provide fundamental insights for the determination of local public health policies, enabling preparedness and consequent mitigation of seasonal rises in incidence.

Several previous studies have taken on the challenge of predicting seasonality of SARS-CoV-2 based on direct analysis of incidence across seasons during the initial pandemic spread ^49–51^. During a zoonotic pandemic, out-of-phase emergence, regional variations in public health intervention, and stochastic pulses of local transmission can obscure the signature of seasonality from surveillance data ^13^. Such concerns have made these analyses controversial ^52,53^. To avoid such concerns, our analyses are based on multi-year endemic coronavirus incidence data and are not subject to the biases introduced by pandemic emergence and large-scale public health interventions. Unlike most other studies, our analysis does not force or even suggest any functional form or *a priori* expectation of seasonality. Instead, our results are driven by incidence data from endemic coronaviruses and informed by their shared evolutionary history. Results from our analysis are broadly consistent with the seasonal incidence trends observed for common human-infecting respiratory viruses in the northern hemisphere ^18^.

Our results on the seasonality of SARS-CoV-2 impart expected incidence trends under endemic conditions. Through two alternative mechanisms, seasonality during its pandemic phase might be either greater or lesser than that expected during subsequent endemicity. On the one hand, the absence of previous exposure and a corresponding naive immune response that are associated with overall higher transmission in a pandemic have the potential to exacerbate peaks and troughs of transmission. In this context, seasonality can be further amplified by an overwhelmed and lagging public health response. As such, we could observe heightened seasonal differences in incidence relative to those seen during endemic spread, overlaid onto peaks and troughs caused by the out-of-phase emergence of pandemic disease ^54^. On the other hand, the mechanisms that are driving the seasonality of coronavirus infections might exert slight influences that are magnified by pathogenic population dynamics year on year ^55^. This resonation to convergence could underlie the observed seasonality of endemic coronaviruses (**Supplementary Appendix Figs. S1–S4**). With a smaller forcing factor that is amplified by pathogen population dynamics, we would expect less seasonality for SARS-CoV-2 during pandemic spread than would be seen in its eventual endemic incidence. In this context, it is possible that not enough time has elapsed for endemic seasonality to be fully realized.

Regardless of how the seasonal dynamics will manifest during this transition from its pandemic phase, our projections provide the expected endemic seasonality.

It is tempting to compare our results to the history of surges throughout the COVID-19 pandemic thus far. For instance, following the initial outbreak, peaks of COVID-19 deaths in Sweden, where interventions were very limited and kept steady, are consistent with our projections of a December-February peak of infection. In much of the rest of the world, however, interventions were more extreme and were unsteadily applied. Relaxation of COVID-19 interventions could explain the recent (Summer 2022) surges of deaths in countries such as Japan or irregular patterns in countries such as Australia or New Zealand. Similar irregular patterns can also be found within countries that had variation in interventions such as vaccine uptake or adherence to public health guidelines, including within the United States ^56,57^. This range of policies and adherence to guidelines confounds direct comparisons ^58^.

The seasonal coronavirus incidences in each location were collected in studies that monitored disease in distinct time spans and that may have been subject to a number of annually varying factors that can drive seasonal trends of respiratory infections. However, in many cases the incidences were obtained across multiple years of sampling. For example, the Stockholm, Sweden dataset ^39^ encompasses 2,093 samples spanning a full decade. Consequently, it is unlikely that the month-to-month average incidences of these long-term datasets are substantially affected by anomalous years. Our results project a seasonal rhythm of SARS-CoV-2 that is broadly similar to the trends observed among many major human-infecting respiratory viruses ^59–61^. This well-known seasonal trend toward greater respiratory incidence in the winter has been ascribed to a number of factors: temperature ^18,62–64^, humidity ^65–68^, solar ultraviolet radiation ^69^, and host behavior ^70^. This trend is typically considered to be muted in the tropics, and reversed in the Southern Hemisphere ^61,71^.

Across the datasets assembled for this study, there was also substantial non-temporal variance in patients that were sampled for coronavirus infection. Some datasets were largely or wholly restricted to infants or children ^41^, whereas others were cross-populational studies aggregating a mix of children, teenagers, and adults ^22,46^. A forecast of absolute case numbers would certainly vary between cohorts ^44^. However, this variance in sampling should not impact our estimates of relative seasonal infection trends. This invariance in seasonal incidence arises because relative incidence in children is strongly correlated with relative incidence in other subsets of the local population ^37^. Any relative scale will work to reveal when higher or lower relative incidence should be expected. Indeed, the relative seasonal patterns for the long-term circulating endemic coronaviruses from our analysis of these datasets are consistent with expectations determined for other seasonal respiratory viruses ^59–61^.

Expanded global surveillance of endemic seasonal coronavirus incidence—especially in the undersampled tropics and Southern Hemisphere—will enhance our understanding of coronavirus seasonality and facilitate preparedness. Denser sampling will enable more precise regional estimates. Sampling in the tropics would enable testing of the muted seasonality that appears there; sampling in the Southern Hemisphere would enable testing of a hypothesis of inverted seasonality compared to the Northern Hemisphere. This information would strengthen the foundation for forecasting not only endemic coronavirus seasonality, but also the seasonality of deadly emergent coronaviruses such as SARS-CoV-2.

Both public health interventions and evolutionary change impact whether the projected seasonality of SARS-CoV-2 will be observed. Transmission could be dampened by the acceleration of vaccination efforts around the world that, like other interventions, have the potential to disrupt erstwhile seasonality. Alternatively, the emergence of novel variants with elevated transmissibility—such as the Delta or Omicron variants ^72–74^—have the potential to thwart public health efforts and impact seasonal trends. Our results suggest that surges of novel COVID-19 variants will frequently coincide with other seasonal endemic respiratory viruses including influenza and respiratory syncytial virus ^75,76^, potentially overwhelming healthcare facilities. Our projections affirm the need for systematic, prescient public health interventions that are cognizant of seasonality.

Foreknowledge of seasonality will enable informed, advanced public health messaging regarding seasons of high concern that could help to overcome barriers of nonadherence. Even with widespread vaccination efforts, SARS-CoV-2 is poised to join HCoV-229E, HCoV-NL63, HCoV-OC43, and HCoV-HKU1 as an endemic coronavirus ^77^. For epidemiological inferences such as seasonality that require long-term datasets, evolutionary biology can provide the theoretical foundation to deliver swift, quantitative, and rigorous insight into how novel threats to human health may behave. Our approach provides guidance for myriad public health decisions until the pandemic phase of SARS-CoV-2 spread has passed and collection of long-term data on endemic COVID-19 incidence becomes feasible. Moreover, in future research it can be broadly applied to seasonal data from any group of viruses to forecast the endemic traits of any emergent threat.

## Data Availability

Data and materials availability
All data, inferred phylogenetic trees, imputed monthly proportions, and code underlying this study are publicly available on Zenodo: DOI:10.5281/zenodo.5274735.

https://zenodo.org/record/5274735#.YfGrBRPMIeY

## Acknowledgments

We thank Dan Warren for helpful discussion at the inception of this work.

## Authors contributions

JPT and AD conceived the project and designed the study; ADL and CN performed literature review with contributions from AD, ADT, and AAN; ADL, CN, and AAN accessed, processed, and curated seasonality data; ADL performed formal analyses with guidance from AD, JPT, and HBH; AD, ADL, and JPT designed and implemented data visualizations; JPT and AD wrote the manuscript; ADL, ADT, PS, and AAN contributed components of the manuscript; and all authors reviewed the manuscript before submission. JPT and AD were responsible for the decision to submit the manuscript. All authors had full access to all the data in the study and had final responsibility for the decision to submit for publication. Data was verified by ADL.

## Data and materials availability

All data, inferred phylogenetic trees, imputed monthly proportions, and code underlying this study are publicly available on Zenodo: DOI:10.5281/zenodo.5274735.

## Supplementary Information

Supplementary Information is available for this paper.

Correspondence and requests for materials should be addressed to Jeffrey P. Townsend.

