## Supplemental materials for "Projecting the seasonality of endemic COVID-19"

### Supplementary materials

**Contents**

Investigators … … … … … … … … … … … … … … … … … … … 02
Supplemental Figure S1 … … … … … … … … … … … … … … … … … … 03
Supplemental Figure S2 … … … … … … … … … … … … … … … … … 04
Supplemental Figure S3 … … … … … … … … … … … … … … … … 05
Supplemental Figure S4 … … … … … … … … … … … … … … … 06
Supplemental Figure S5 … … … … … … … … … … … … … … … … 07
Supplemental Figure S6 … … … … … … … … … … … … … … … … … 08
Supplemental Figure S7 … … … … … … … … … … … … … … … … … … 09
Supplemental Figure S8 … … … … … … … … … … … … … … … … … … … 10

**Investigators**
Jeffrey P. Townsend[1,2,3,4*],
April D. Lamb[5],
Hayley B. Hassler[1],
Pratha Sah[6],
Aia Alvarez Nishio[7],
Cameron Nguyen[5],
Alexandra Tew[5],
Alison P. Galvani[6],
Alex Dornburg[5]

**Affiliations**
[1]*Department of Biostatistics, Yale School of Public Health; New Haven, Connecticut, USA*
[2]*Department of Ecology and Evolutionary Biology, Yale University; New Haven, Connecticut, USA*
[3]*Program in Computational Biology and Bioinformatics, Yale University; New Haven, Connecticut, USA*
[4]*Program in Microbiology, Yale University; New Haven, Connecticut, USA*
[5]*Department of Bioinformatics and Genomics, University of North Carolina at Charlotte; Charlotte, NC*
[6]*Center for Infectious Disease Modeling and Analysis, Yale University; New Haven, Connecticut 06510, USA*
[7]*Yale College; New Haven, Connecticut 06510, USA*

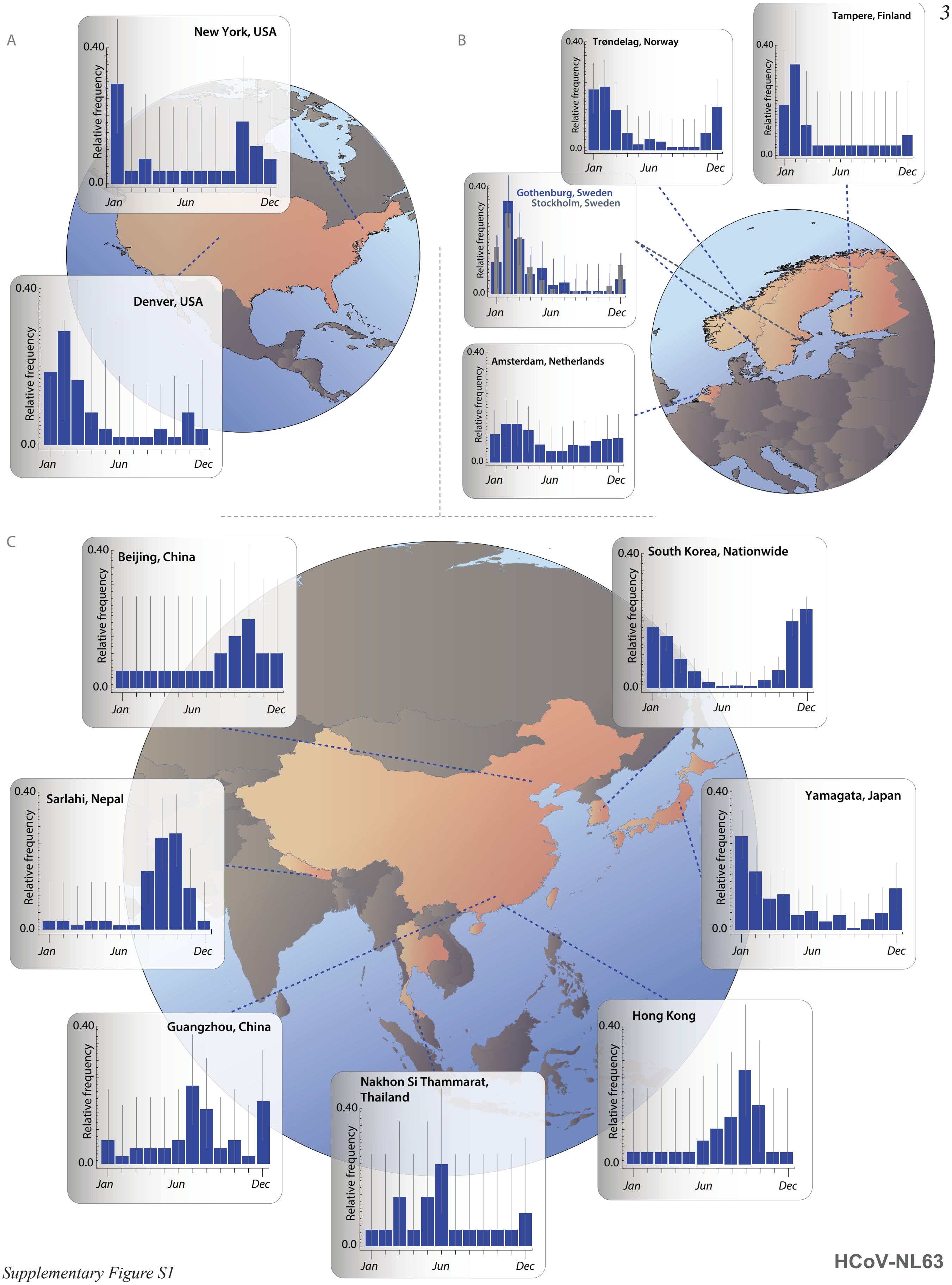


*Supplementary Figure S1*

Proportions of HCoV-NL63 infections by month across locations in (A) North America, (B) Europe, and (C) Asia. Vertical gray bars indicate 95% confidence interval of raw monthly infections.

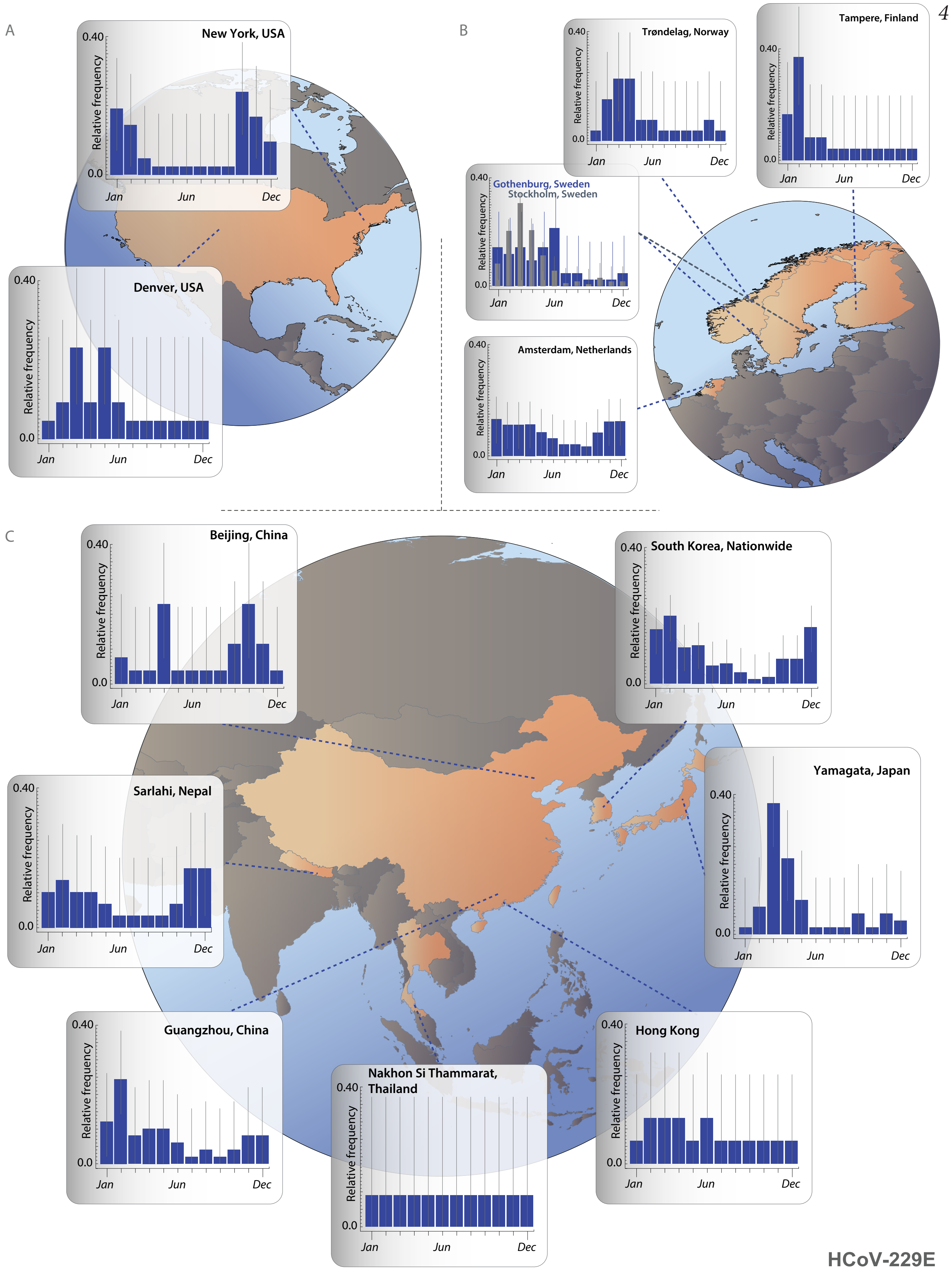


*Supplementary Figure S2*

Proportions of HCoV-229E infections by month across locations in (A) North America, (B) Europe, and (C) Asia. Vertical gray bars indicate 95% confidence interval of raw monthly infections.

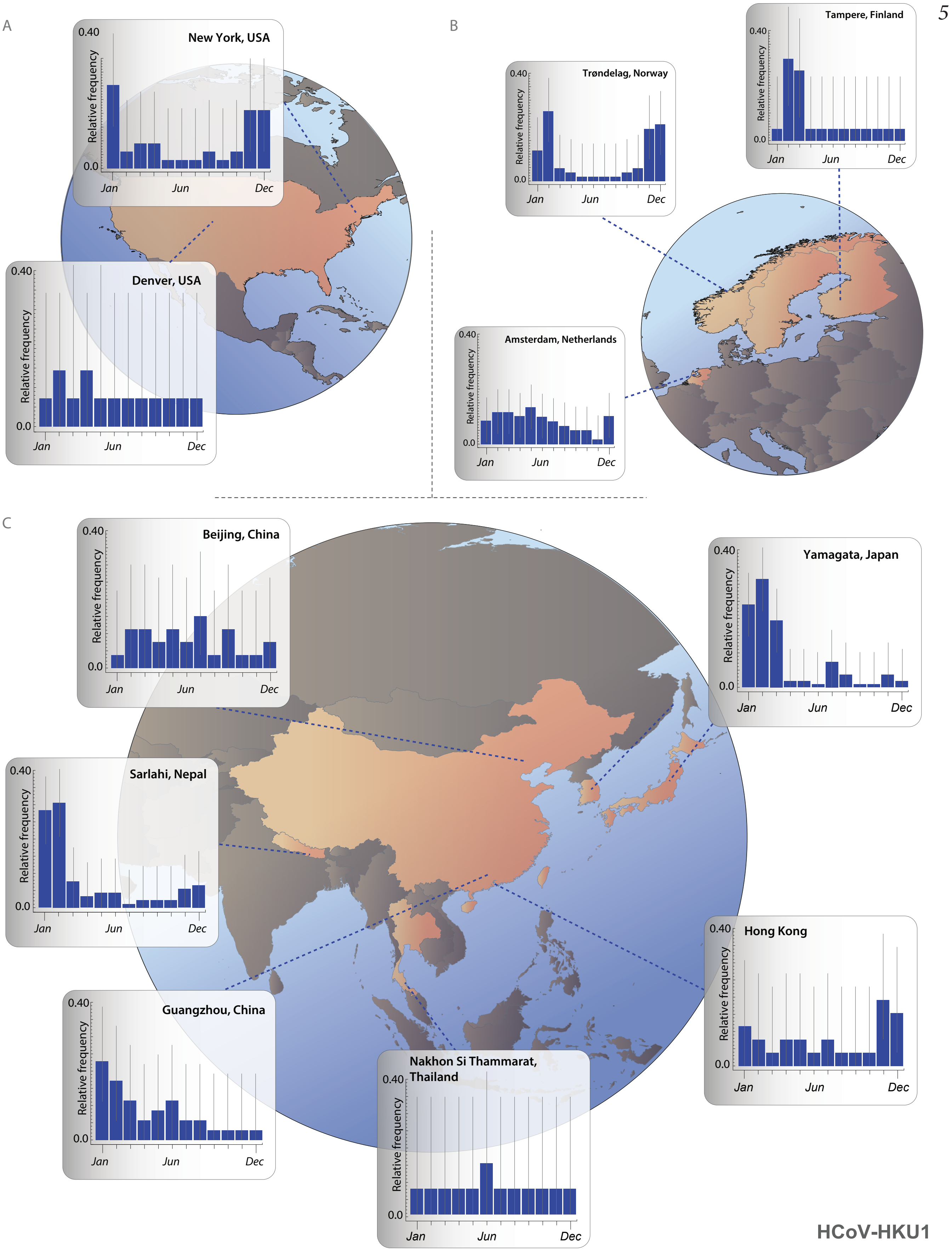


*Supplementary Figure S3*

Proportions of HCoV-HKU1 infections by month across locations in (A) North America, (B) Europe, and (C) Asia. Vertical gray bars indicate 95% confidence interval of raw monthly infections.

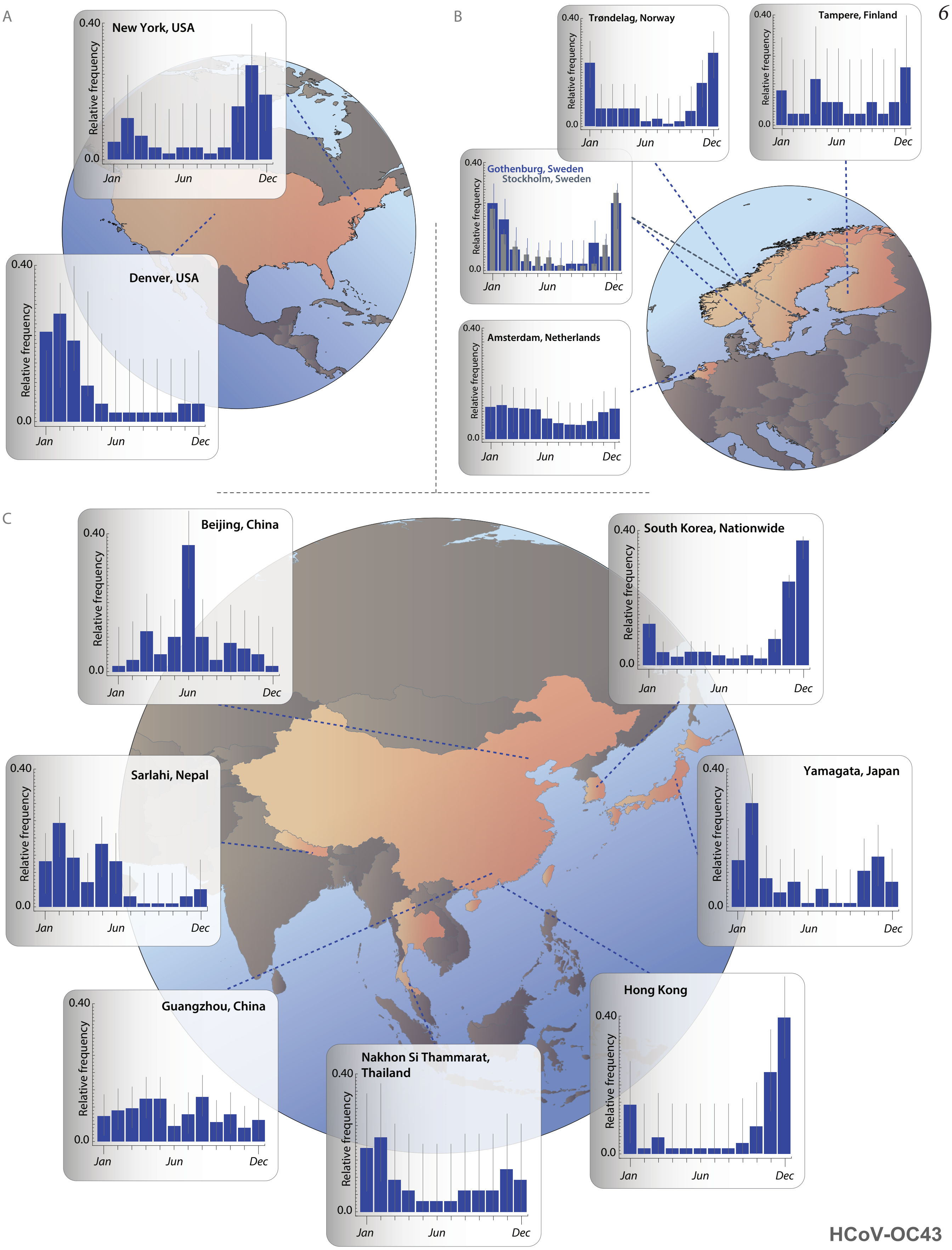


*Supplementary Figure S4*

Proportions of HCoV-OC43 infections by month across locations in (A) North America, (B) Europe, and (C) Asia. Vertical gray bars indicate 95% confidence interval of raw monthly infections.

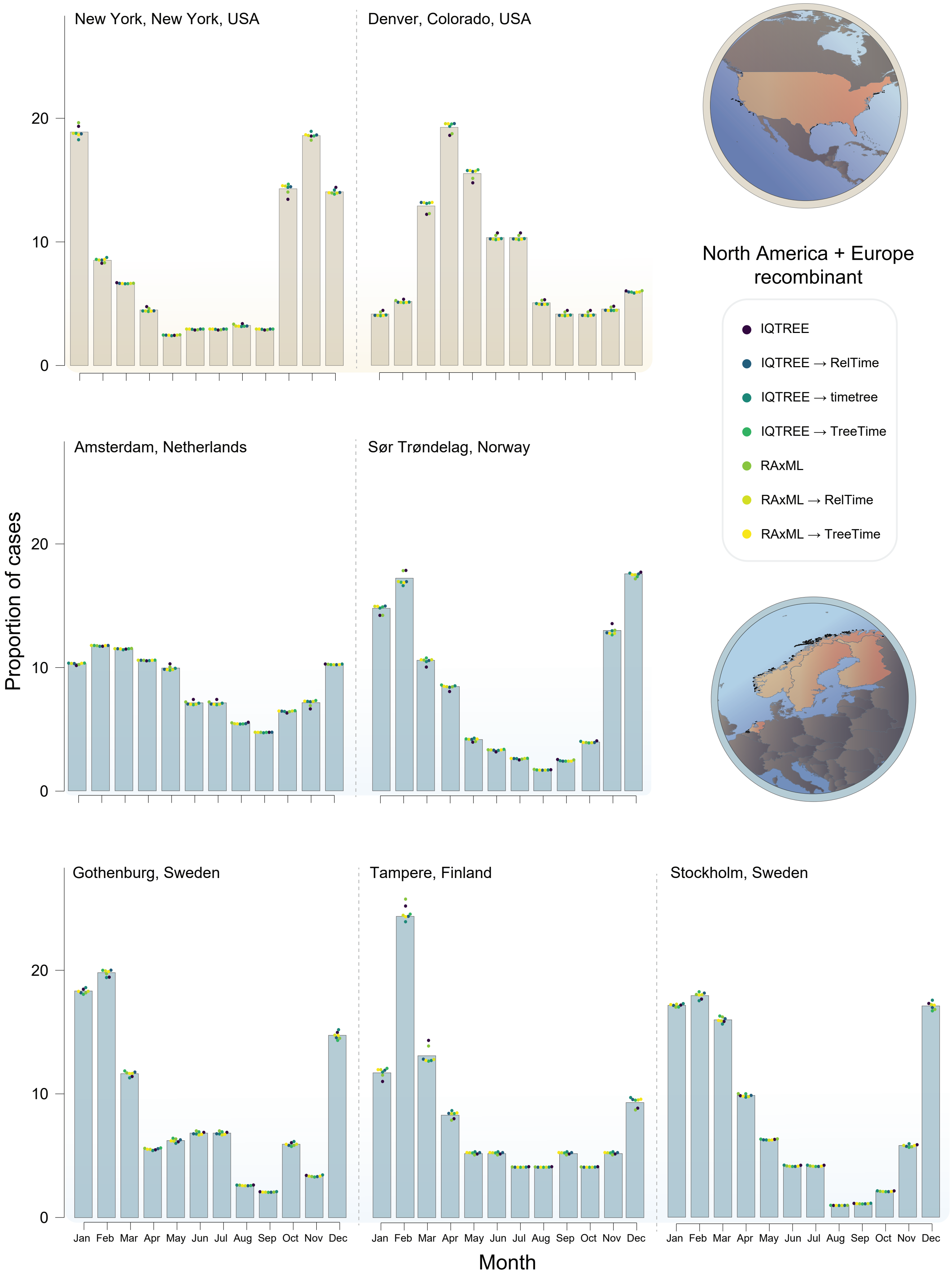


*Supplementary Figure S5*

Sensitivity of proportions of SARS-CoV-2 infections by month to method of time tree inference in North America (tan) and Europe (blue).

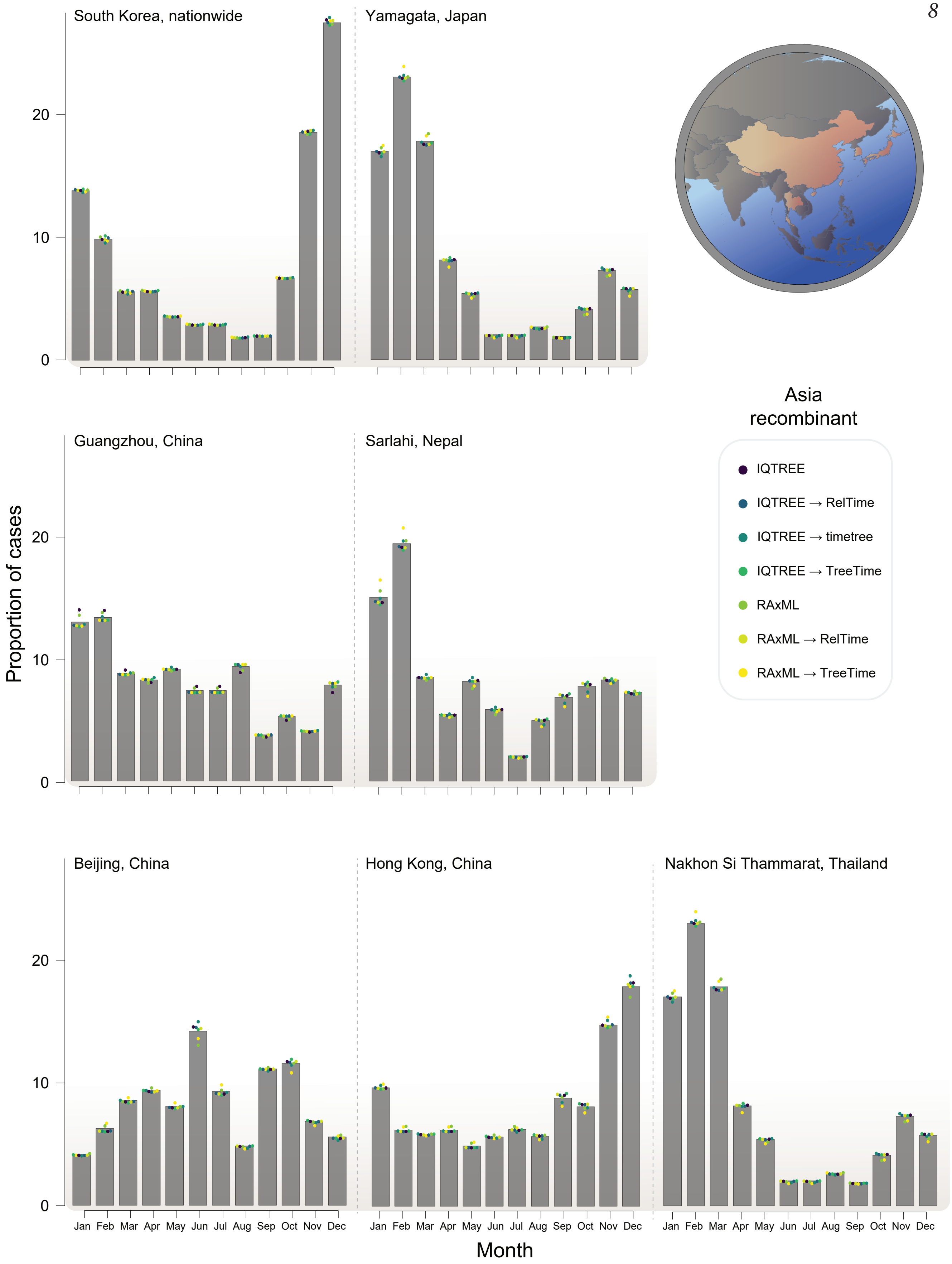


*Supplementary Figure S6*

Sensitivity of proportions of SARS-CoV-2 infections by month to method of time tree inference in Asia.

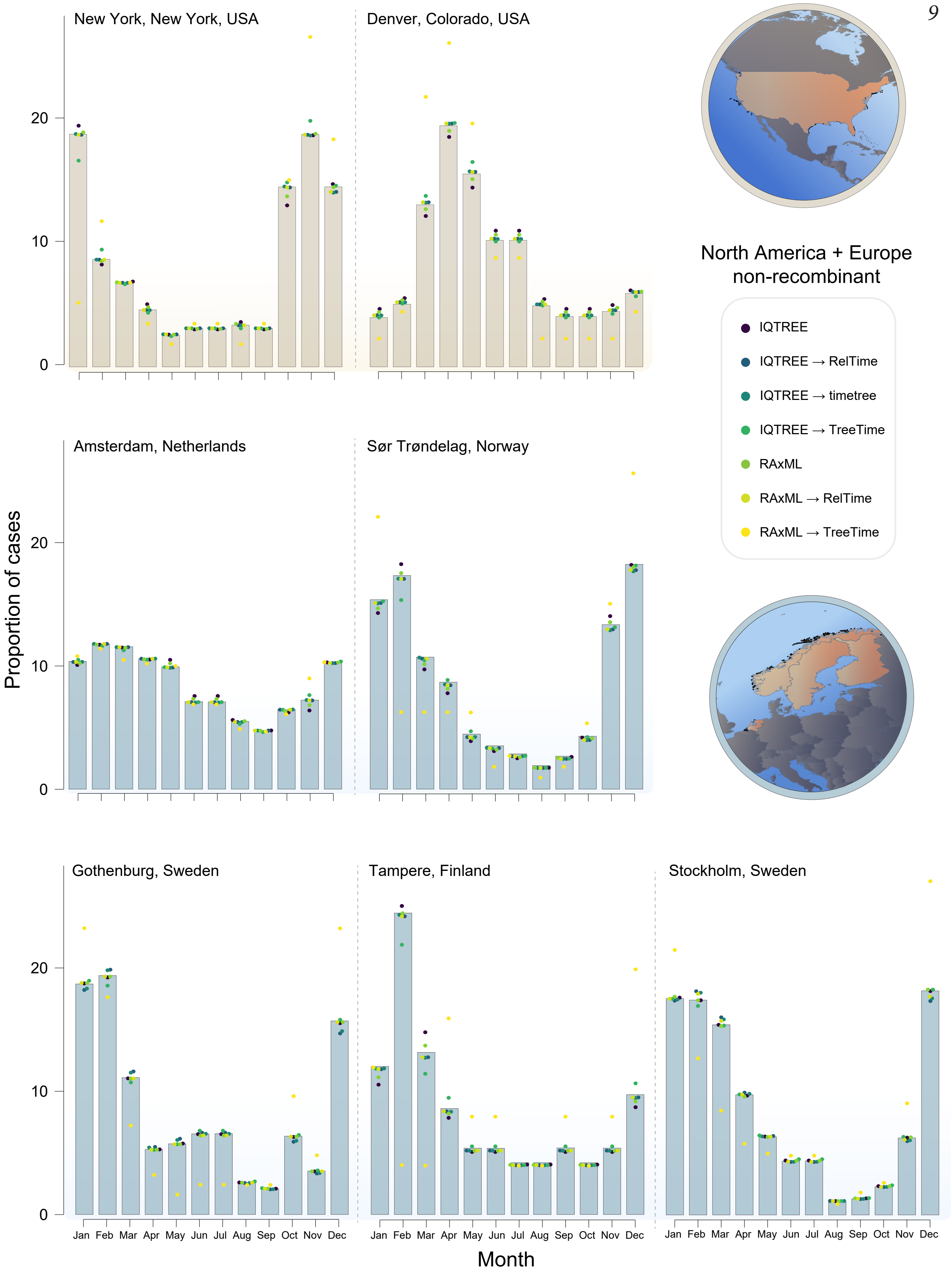


*Supplementary Figure S7*

Sensitivity of proportions of SARS-CoV-2 infections by month to use of only viral sequences deemed non-recombining by location, in North America (tan) and Europe (blue).

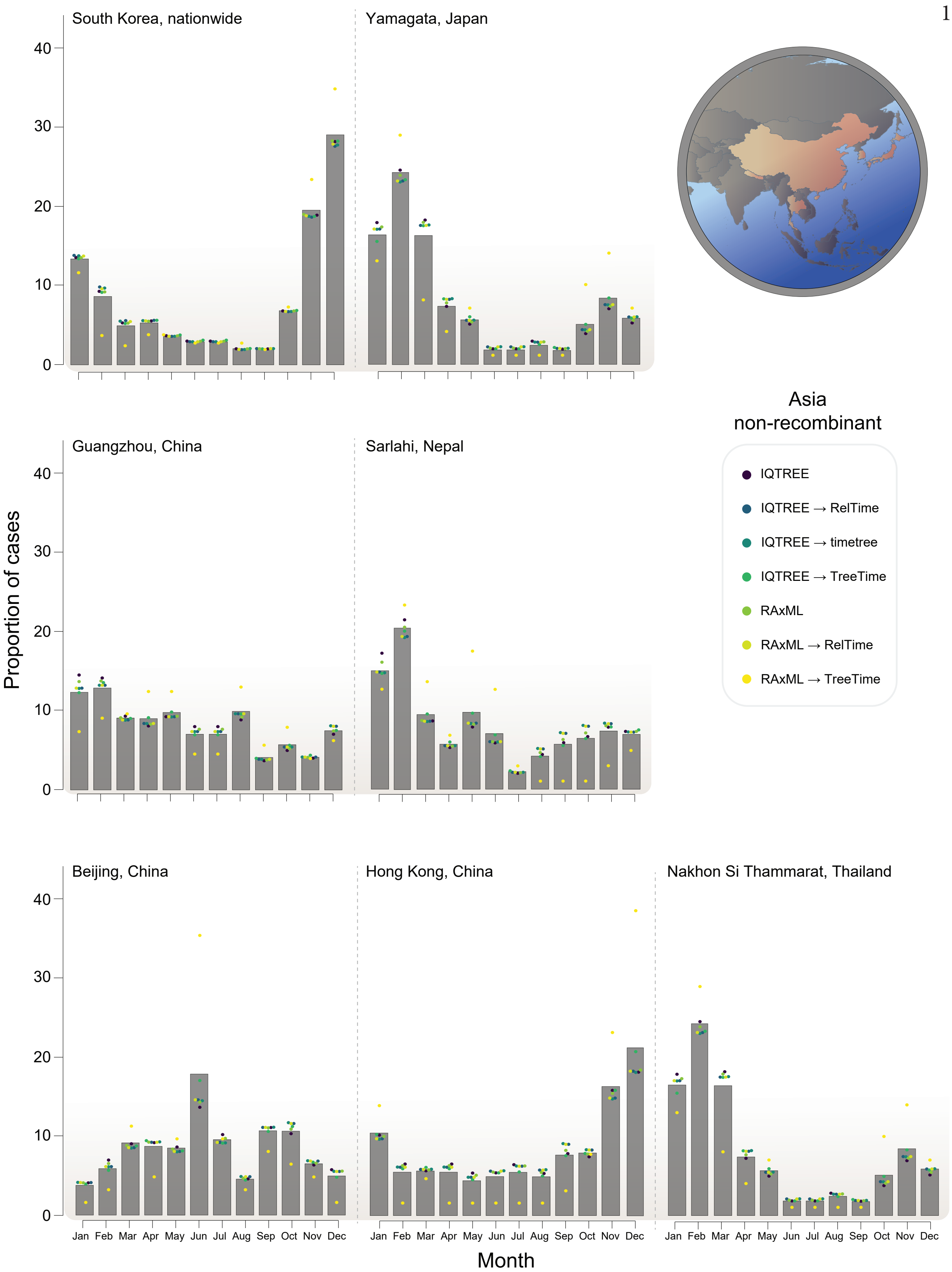


*Supplementary Figure S8*

Sensitivity of proportions of SARS-CoV-2 infections by month to use of only viral sequences deemed non-recombining by location, in Asia.
